# The Relationship between Weekly Periodicity and COVID-19 Progression

**DOI:** 10.1101/2020.11.24.20238295

**Authors:** Sophia Li

**Affiliations:** Massachusetts Institute of Technology

**Keywords:** COVID-19 progression, weekly periodicity, strength and persistence indicators, societal and institutional responses, correlation to pandemic outcome

## Abstract

COVID-19 is extraordinary both as once-in-a-lifetime pandemic and having abundant real-time case data, thus providing an extraordinary opportunity for timely independent analysis and novel perspectives. We investigate the weekly periodicity in the daily reported new cases and new deaths with the implied relationships to the societal and institutional responses using autocorrelation and Fourier transformation. The results show significant linear correlations between the weekly periodicity and the total cases and deaths, ranging from 50% to 84% for sizable groups of countries with population normalized deaths spanning nearly three orders of magnitude, from a few to approaching a thousand per million. In particular, the Strength Indicator of the periodicity in the new cases, defined by the autocorrelation with a 7-day lag, is positively correlated strongly to the total deaths per million in respective countries. The Persistence Indicator of the periodicity, defined as the average of three autocorrelations with 7-, 14- and 21-day lags, is an overall better indicator of the progression of the pandemic. For longer time series, Fourier transformation gives similar results. This analysis begins to fill the gap in modeling and simulation of epidemics with the inclusion of high frequency modulations, in this case most likely from human behaviors and institutional practices, and reveals that they can be highly correlated to the magnitude and duration of the pandemic. The results show that there is significant need to understand the causes and effects of the periodicity and its relationship to the progression and outcome of the pandemic, and how we could adapt our strategies and implementations to reduce the extent of the impact of COVID-19.

## 1 Introduction

One of the defining events of 2020 and perhaps this century is COVID-19, a pandemic that has swept through 213 countries, totaling 29 million cases and claiming 930 thousand deaths as of September 13th[1]. With the pandemic lasting for over nine months now, long term health effects unknown, and the second wave of infections looming as the Northern Hemisphere enters the fall season, the urgent need to find all the causes underlying the relentless progression of COVID-19 is greater than ever to prevent more infections and deaths.

As many governments and organizations still try to choose between disease containment and economic impact of COVID-19, it is important to recognize that pandemic deaths and economic contractions are closely linked - prolonged shutdowns and restrictions due to the continuing growth of COVID-19 cases and deaths lead to contracting economies. Reopening of societies and recovering of economies are hampered severely by the ongoing pandemic. However it does not take a prolonged lockdown to control and mitigate if the key aspects of the pandemic progression are recognized, and corresponding measures designed and utilized[2, 3]. To prevent the economy from further contractions due to COVID-19 or new disruptions by the incoming second or third wave, it would be prudent to prevent further infections and deaths by better modeling what approaches could help with the broadest practical feasibility now, and contain the pandemic sooner.

A unique characteristic of COVID-19 pandemic is the abundance of open real-time case data worldwide. Most governments are reporting the number of new cases and new deaths every day, allowing novel approaches to analyze the data, and creating a unique opportunity for understanding more aspects of the hidden relationships and underlying dynamics of the progression of such infectious diseases.

The World Health Organization and most governments realized the dire threat of this pandemic early on, and declared states of national emergency and instituted a variety of control and containment policies. Different countries have reacted rather differently to the pandemic, differing in policies and reactions from citizens and institutions. Looking at the total deaths per million and the total cases per million, it is noticeable that the population normalized total deaths differ up to three orders of magnitude for all countries and regions, ranging from fewer than 1 in a million to close to 1000 in a million [4].

Besides the vast differences in outcomes, another striking pattern is seen in the daily reported data for new cases and new deaths. A regular modulation is observed, as shown in Figure 1 below. Such modulation is found in many countries, differing in strength and persistence, and may offer additional insight into how different policies and practices affect the containment of this infectious disease. A 7-day periodicity is not a new observation. The trend is fairly obvious and can be visually identified with some simple treatment of the data[5, 6], and becomes even more explicit when one performs a Fourier transformation to find peaks at 1/7 and 1/14 frequencies from sufficiently long time series[7].

**Figure 1:**
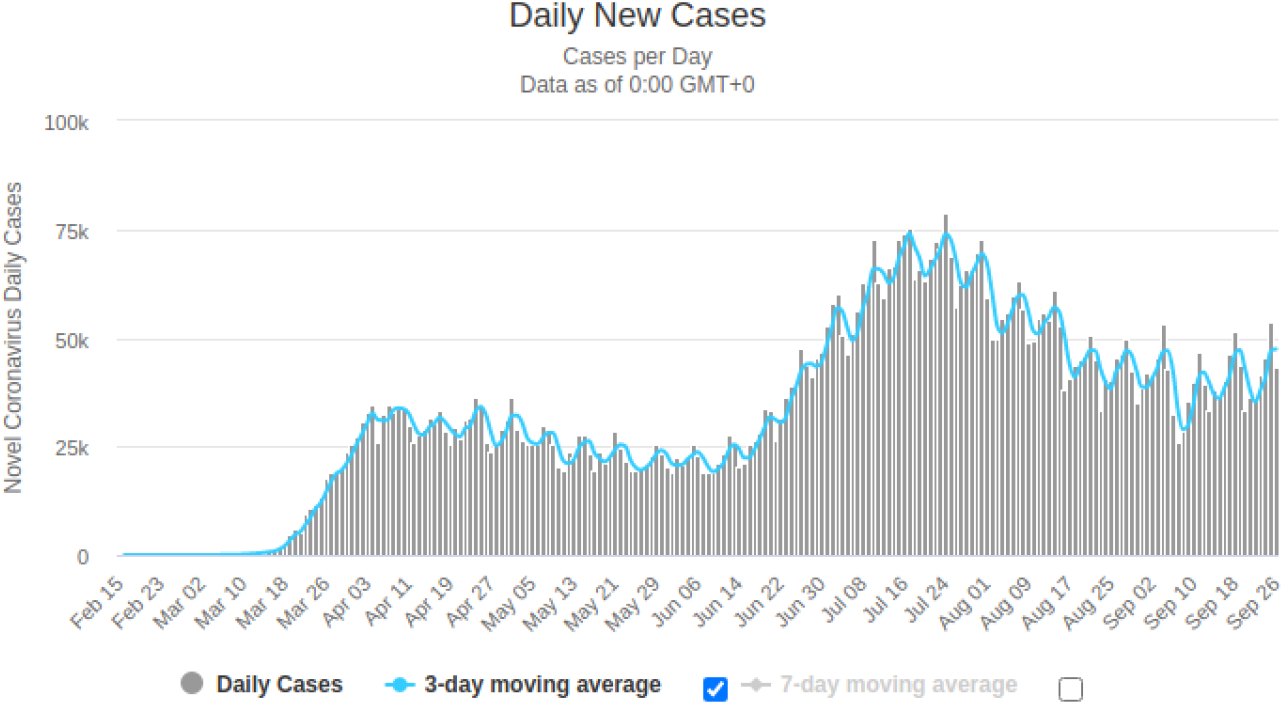
Raw data of the daily new cases in USA from Worldometers[19].

To predict and help curb the number of cases and deaths, many models specific to COVID-19 have been developed to better recognize causes for the growth and spread of the disease, predict the potential outcomes, or create planning scenarios for the pandemic responses. Models have been developed to consider more variables, such as deaths and unreported cases, for parameter calibration of their models[8, 9, 10, 11], taking into account a latency period[12, 13], and looking at non-pharmacological parameters like human behavior or socioeconomic background[14, 15]. These models and simulations are all designed to better predict the pandemic progression and potential outcomes. Most offer insights and ideas on how to better control and contain COVID-19. However, none of these models uses high-frequency periodicity as a parameter or indicator of the pandemic progression. In fact, short-term modulations in the data are commonly averaged out and not included in the equations and models to show the broader and longer-term patterns and results. The prior studies of periodicity in epidemics often are highly theoretical, and typically look at low-frequency periodicity over seasons or years, and generally are used to predict the next waves of an epidemic[16, 17]. In particular, studies on low-frequency periodicity in COVID-19 project wintertime outbreaks four to five years after the initial outbreak[18]. These studies neglect the effects that high-frequency modulations for high-growth infections can have on the magnitude and duration of the low-frequency waves, and provide little guidance on the ongoing responses to rapidly progressing pandemics.

The existence of a high-frequency modulation pattern in the daily new cases and deaths of COVID-19 has not been explored systematically and may provide another perspective to discover hidden factors affecting the levels of containment. In [7], the 7-day periodicity was confirmed by performing a Fourier transformation on the raw data for select countries, but the analysis did not further explore the meanings of the signals. It only touched on the influence human and cultural habits have on the disease spread. On the surface, it would seem highly unlikely that short-term fluctuations in the case data, regularly compiled and reported after the fact, could be related to the outcome of the pandemic. In this study, we attempt to connect the periodicity signal to the progression and outcome quantitatively and systematically, and make this periodicity an indicator of the effectiveness of containment responses.

There are three necessary parts for infections to spread: a source, susceptible hosts, and a transmission path[20]. The source of COVID-19 is a novel coronavirus SARS-CoV-2. Although the number of infected is high, the total cases only represent fewer than 1% of the world population, so almost everyone is still a susceptible host and may become a carrier and source. The development of vaccines to immunize hosts and minimize sources is highly technical and ongoing, and their wide uses are months away at least, whereas the long path to herd immunity to decrease susceptible hosts will cost many more human lives given the relatively high case fatality rates. Some effective intermediate approaches to control and contain the pandemic are essential.

Like all the studies conducted and models created to explain and predict COVID-19 spread, the goal of this study is to provide proof on one of the possible causes and consequences of pandemic progression by investigating certain hidden relationships, that may potentially influence policy making, human behaviors, social interactions, and business practices to more effectively control and minimize the long term damages caused by COVID-19.

The scope of this study will focus on the second and third aspects of infectious spread, susceptible hosts and transmission paths. The observed periodicity is unlikely due to the virus itself since the reported data from many countries do not show such modulations, and is much more likely to be related to the availability of hosts and the paths. The findings in the study suggest that the differences in personal and institutional responses may have contributed substantially to the large differences in periodicity, and to change the availability of hosts and paths will require changes in our individual and collective behaviours during the pandemic. We should be able to modify how we work and live temporarily during the most critical periods, disrupt the routines that may inadvertently enhance the availability of hosts and means, and improve the eventual outcome at minimal total costs to safety and economy, as some countries have demonstrated.

## 2 Methods

We performed autocorrelation computation and Fourier transformation on the data for the daily new cases per million and the new deaths per million for countries with statistically meaningful data. We correlated the strength (7-day lag peak heights) and persistence (average of three peak heights, 7-, 14-, and 21-day lags) of the autocorrelation and Fourier transformation signals to the total deaths per million and the total cases per million as new indicators of disease containment. All computations and analyses were performed with Python, using tools of SciPy [21], NumPy [22], and Matplotlib [23].

### 2.1 Data Treatment

The data comes from Our World in Data [4]. We removed countries [Table 1] that did not have statistically meaningful numbers of new cases or new deaths due to COVID-19 needed to produce clear signals. Some of these countries, such as Thailand and New Zealand, treated and contained COVID-19 in a timely manner, whereas some others are not actively reporting COVID-19 cases, likely indicating they do not have a problem. Others, like San Marino or the Vatican are excluded from the study due to their very small population sizes. We also removed the outlying data points representing very unique situations - Belgium has an abnormally high count of total deaths per million, probably due to an aging population extremely susceptible to death during the initial encounter with insufficient awareness and preparedness; countries like Qatar and Bahrain have unusually large numbers of total cases per million because they test their populations at much higher rates than most other countries[24]. We will refer to the remaining 149 countries as “all countries” in the rest of this analysis.

**Table 1:** The countries are grouped into these three categories. Removed countries are not analyzed in this study for lack of statistically significant case data. “First wave” countries have 10 cases per million by March 20th, while “second wave” countries are all the remainders.

| Removed Countries | “First Wave” Countries | “Second Wave” Countries |
| --- | --- | --- |
| Andorra, Angola, Anguilla, Aruba, Bahrain, Barbados, Belize, Belgium, Bhutan, Bonaire Sint Eustatius and Saba, Botswana, British Virgin Islands, Burundi, Cambodia, Cayman Islands, Cote d’Ivoire, Curacao, Democratic Republic of Congo, Dominica, El Salvador, Eritrea, Faeroe Islands, Falkland Islands, Fiji, French Polynesia, Gibraltar, Greenland, Grenada, Guam, Guinea, Hong Kong, Jersey, Laos, Lebanon, Lesotho, Mauritius, Mongolia, Montserrat, Nepal, New Caledonia, New Zealand, Niger, Northern Mariana Islands, Papua New Guinea, Rwanda, Saint Kitts and Nevis, Saint Lucia, Saint Vincent and the Grenadines, San Marino, Sao Tome and Principe, Seychelles, Sierra Leone, Timor, Turks and Caicos Islands, Uganda, United Arab Emirates, Vatican, Vietnam, Western Sahara, Yemen | Austria, Brunei, Burkina Faso, China, Cuba, Denmark, Estonia, France, Georgia, Germany, Iceland, Indonesia, Iran, Italy, Jordan, Liechtenstein, Luxembourg, Malaysia, Malta, Monaco, Netherlands, Nigeria, Norway, Qatar, Senegal, Slovenia, South Korea, Spain, Sri Lanka, Sudan, Sweden, Switzerland, Taiwan, Thailand, Togo, Uruguay | Afghanistan, Albania, Algeria, Antigua and Barbuda, Argentina, Armenia, Australia, Azerbaijan, Bahamas, Bangladesh, Belarus, Benin, Bermuda, Bolivia, Bosnia and Herzegovina, Brazil, Bulgaria, Cameroon, Canada, Cape Verde, Central African Republic, Chad, Chile, Colombia, Comoros, Congo, Costa Rica, Croatia, Cyprus, Czech Republic, Djibouti, Dominican Republic, Ecuador, Egypt, Equatorial Guinea, Ethiopia, Finland, Gabon, Gambia, Ghana, Greece, Guatemala, Guernsey, Guinea-Bissau, Guyana, Haiti, Honduras, Hungary, India, Iraq, Ireland, Isle of Man, Israel, Jamaica, Japan, Kazakhstan, Kenya, Kuwait, Kyrgyzstan, Latvia, Liberia, Libya, Lithuania, Macedonia, Madagascar, Malawi, Maldives, Mali, Mauritania, Mexico, Moldova, Montenegro, Morocco, Mozambique, Myanmar, Namibia, Nicaragua, Oman, Pakistan, Palestine, Panama, Paraguay, Peru, Philippines, Poland, Portugal, Puerto Rico, Romania, Russia, Saudi Arabia, Serbia, Singapore, Sint Maarten (Dutch part), Slovakia, Somalia, South Africa, South Sudan, Suriname, Swaziland, Syria, Tajikistan, Tanzania, Trinidad and Tobago, Tunisia, Turkey, Ukraine, United Kingdom, United States, United States Virgin Islands, Uzbekistan, Venezuela, Zambia, Zimbabwe |

**Table 2:** These countries have either a large number of total deaths, or strong weekly periodicity evidenced by autocorrelation.

| “Filtered Countries” |
| --- |
| Armenia, Austria, Bangladesh, Brazil, Bulgaria, Central African Republic, Chad, Chile, Croatia, Cyprus, Czech Republic, Denmark, Dominican Republic, Ecuador, Estonia, Finland, France, Gabon, Gambia, Germany, Guatemala, Guernsey, India, Indonesia, Israel, Italy, Kazakhstan, Kuwait, Luxembourg, Macedonia, Mexico, Moldova, Netherlands, Norway, Panama, Paraguay, Philippines, Poland, Portugal, Romania, Senegal, Slovakia, Slovenia, Somalia, South Africa, Spain, Sweden, Switzerland, United Kingdom, United States, Zambia |

As reporting mechanisms and standards are evolving while we learn more about the disease, there are often discrepancies or changes in the raw data. Negative numbers indicate that a country has over-reported previously, while outlying large spikes indicate that a country has under-counted or under-reported. Since we are interested in periodic patterns, unusual data of this kind will not alter the periodicity in significant ways in the analysis. Therefore the negative numbers are replaced with zero and the outlying numbers, found using the built-in SciPy tool *find peaks*, are normalized with the average value of the data the days before and after the day of the outlying data.

These replacements are reasonable and sufficient for the present analysis; improved data treatment can be performed later. As a possible way of normalizing the data, the unusual data can be spread or taken out proportionally over the prior data points. Such treatment would only change the amplitude but not the underlying periodicity. In a hypothetical situation where we spread out the unusual data to the previously reported data, we would see that the amplitude of the prior data would be changed from 20% to 22% if the average prior data amplitude was 20% and the unusual data represented a 10% increase in all the prior data. This would not significantly change the peak amplitude of the computational output of autocorrelation and Fourier transformation, especially since the periodicity has not changed. Finally, to remove the overarching trend over weeks and months in the weekly periodicity analysis, the treated raw data is subtracted with a centered seven-day moving average.

Even after removing many countries (60) from the analysis, there remains a large number countries (149) to be analyzed. Categorizing countries further can give more meaning to the analysis, as well as make the results more specific. Most studies categorize countries by geographical regions or income levels, but since this study is focused on exploring the reasons behind global trends with universal indicators, countries are separated by when they first reach a critical amount of total cases. Countries that have more than 10 cases per million before March 20th are considered “first wave” countries (36) in this study, while all other countries (113) are “second wave” countries. It is important to note that the uses of “first wave” and “second wave” here indicate the chronological orders of encountering substantial infections, and are different from the terminology used by healthcare professionals for large scale recurring infections. Separating the countries using the criteria described above, we can analyze how countries reacted depending on the information available to them at the moment, and the corresponding progression and outcomes [Table 1].

**Figure 2:**
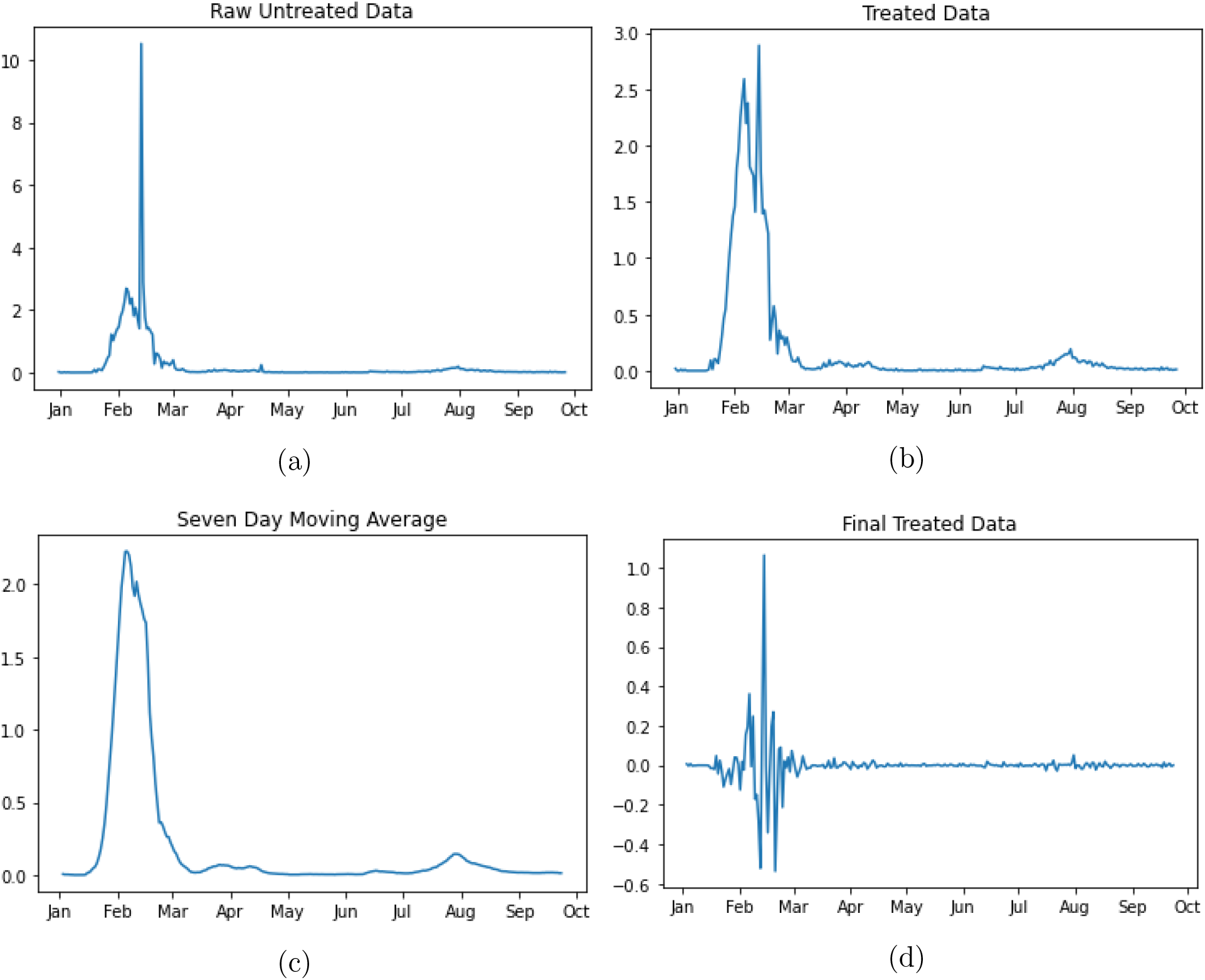
This is a graphic example of how the data is treated in this study. 2a is the raw data, and 2b shows the data after removing the outlying unusual point from the raw data. 2c is the 7-day moving average using 2b. By subtracting 2b with 2c, we obtain 2d used in the following analysis.

### 2.2 Analysis Methods

Autocorrelation reveals the correlation between the values of a data set across different time lags in the observations[25]. Autocorrelation compare the similarity of the data to a copy of itself that has a time delay. It is a useful tool to find patterns or prove randomness. We use autocorrelation to prove there is a 7-day repetitive pattern, and find how strong and persistent it is. Using Python to compute the autocorrelation for each country, we quantify the results using two methods to create two different autocorrelation indicators. The first indicator uses the peak value from the 7-day lag, which primarily measures the strength of the periodicity; the second uses the average from the 7-, 14-, and 21-day lags, which indicates the persistence of the periodicity, looking at the continued strength of the signals for lags up to three weeks. To prevent potential confusion with the (cross-)correlations below, we will call the amplitude of the first autocorrelation peak the Strength Indicator, and the averages of the three peaks the Persistence Indicator. These autocorrelation indicators, or Strength and Persistence, are plotted separately against the total death per million and total case per million in a scatter plot, a linear regression is performed, and a coefficient of determination (r-squared) or a coefficient of correlation (r) is found. We use the total death per million as a measure of disease containment and patient care, and total case per million as an indicator for the societal response to the disease. Since death is the final and irreversible loss of human life, it is a stronger and more important indicator of the outcome.

**Figure 3:**
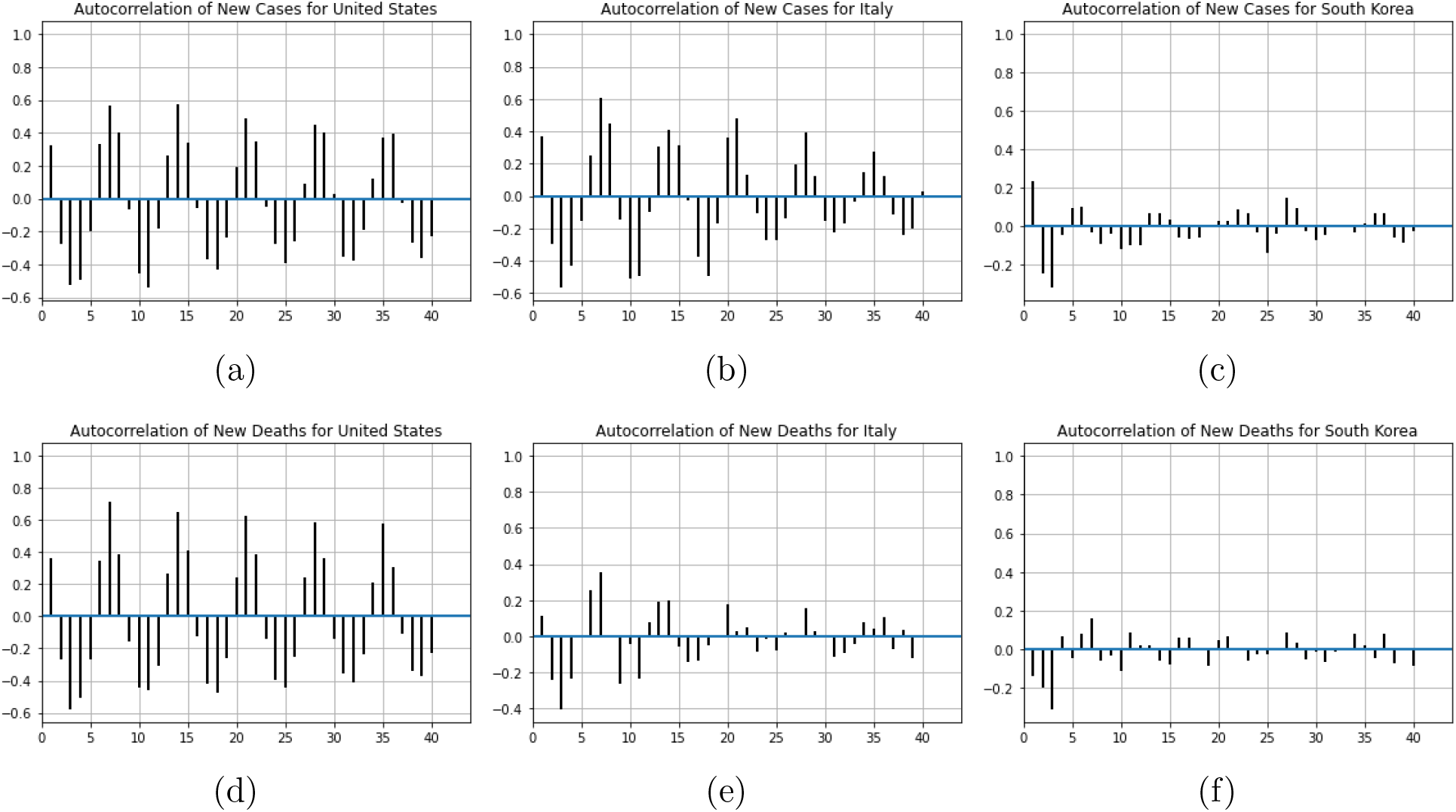
Examples of autocorrelation results. 3a and 3d show strong and persistent autocorrelation. 3b shows strong but declining autocorrelation. 3c, 3e, and 3f show weak autocorrelation.

In addition to the above analysis, we select the countries with data significant in the following manner: countries with a first autocorrelation peak, the Strength Indicator, higher than 0.2, or having more than 10 total cases per million, and perform a similar analysis with these countries. We will call these countries the “filtered countries”, since their case data show very clear periodicity, and may reveal more distinct relationships.

A Fourier transformation decomposes the data into it’s constituent frequencies, and we can use it to find significant frequencies in the data. Due to the fact that the data is discrete because new cases and new deaths are reported daily and therefore individual and separate, we can conduct a discrete Fourier analysis. We choose to perform a fast Fourier transformation, which is an optimized algorithmic method that finds the periodic patterns in the normalized and discrete data, using the Python library SciPy. We find the amplitude of the signal where the frequency is 1/7, and plot that against the total deaths per million and total cases per million 4. This part of the analysis is not separated into two waves. Once again, linear regression is performed to find the coefficient of determination (r-squared) and coefficient of correlation (r).

**Figure 4:**
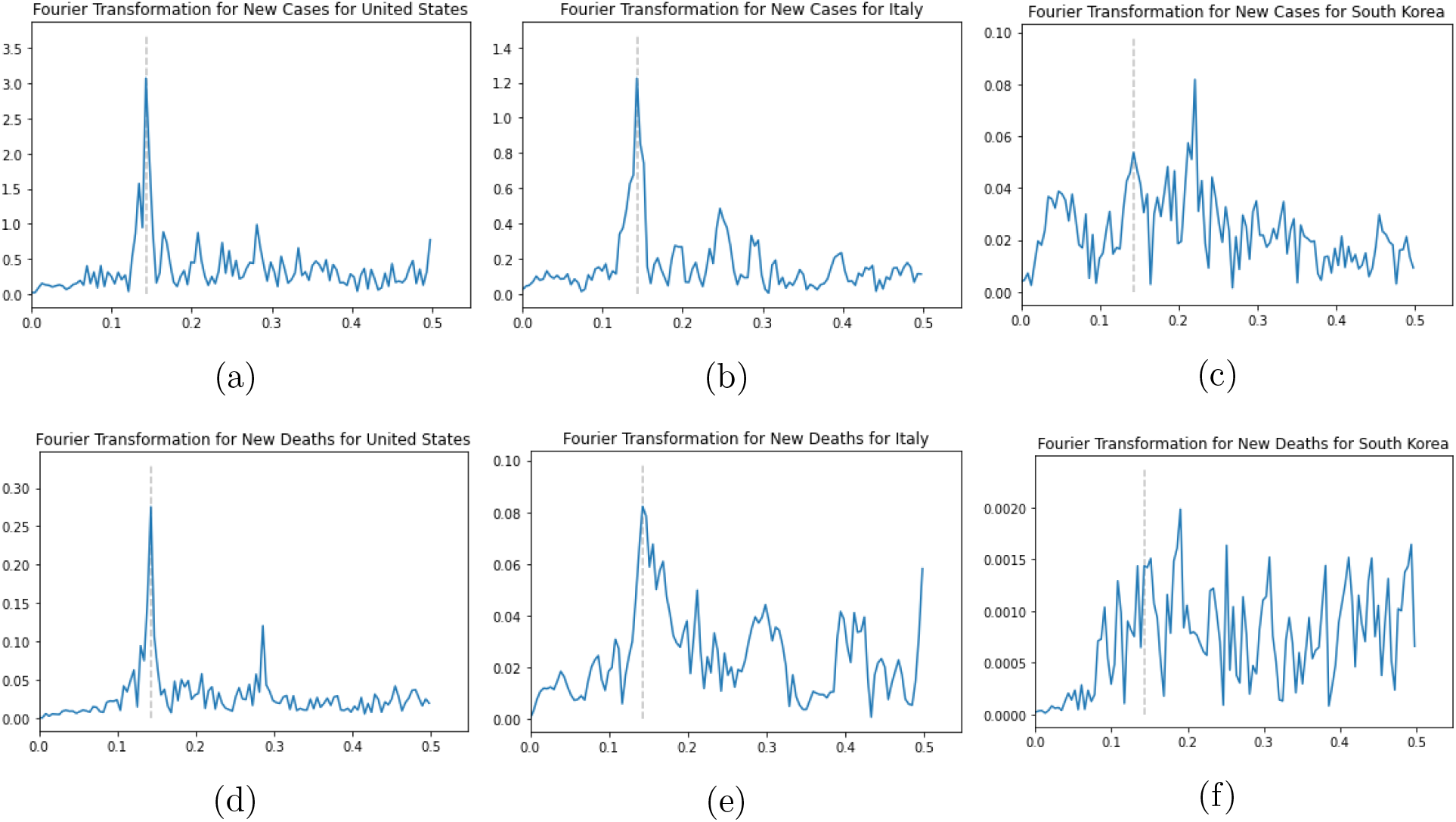
Examples of Fourier transformation results. The dotted line indicates the position of 1/7 frequency. These correspond well with our autocorrelation analysis. Note the differences in the vertical scale.

For this study, the focus is placed more heavily on the autocorrelation than the Fourier transformation because the former requires fewer data points to produce meaningful signals. When starting this analysis in June, the data set was smaller and Fourier transformation gave much poorer results compared to the autocorrelation. Requiring fewer data means that the analysis can be performed earlier in a more timely manner. In the case of a rapidly escalating pandemic, it is important to quickly understand as much as possible about the disease spread so we can improve our responses in real time for faster and better control and containment.

## 3 Results

Within this study, the following tables below display the results [Table 3, 4, 5]. Only the scatter plots for the “all countries” data are presented here, and additional plots for subcategories can be found in the supplementary material section.

**Table 3:** The coefficient of determination and the slopes from these relationships for “all countries” are tabulated here.

| “All Country” Indicator and Results |  |  |  |  |  |  |  |  |  |  |  |  |
| --- | --- | --- | --- | --- | --- | --- | --- | --- | --- | --- | --- | --- |
|  | “All Countries” |  |  |  | “First Wave” |  |  |  | “Second Wave” |  |  |  |
|  | Total Cases per Million |  | Total Deaths per Million |  | Total Cases per Million |  | Total Deaths per Million |  | Total Cases per Million |  | Total Deaths per Million |  |
|  | New Cases | New Deaths | New Cases | New Deaths | New Cases | New Deaths | New Cases | New Deaths | New Cases | New Deaths | New Cases | New Deaths |
| Strength Indicator $R^2$ | 0.125 | 0.066 | 0.244 | 0.25 | 0.292 | 0.091 | 0.487 | 0.394 | 0.115 | 0.076 | 0.172 | 0.199 |
| Strength Indicator Slope | 6913 | 5875 | 342 | 403 | 6526 | 3950 | 531 | 517 | 7117 | 6941 | 276 | 355 |
| Persistence Indicator $R^2$ | 0.155 | 0.107 | 0.309 | 0.329 | 0.292 | 0.198 | 0.567 | 0.433 | 0.154 | 0.110 | 0.233 | 0.293 |
| Persistence Indicator Slope | 10916 | 10402 | 543 | 644 | 9501 | 8583 | 834 | 790 | 11531 | 11465 | 450 | 591 |

**Table 4:** The correlation coefficients from these relationships for “filtered countries” are tabulated here.

| “Filtered Countries” Indicators and Relationships |  |  |  |  |  |  |  |  |  |  |  |  |
| --- | --- | --- | --- | --- | --- | --- | --- | --- | --- | --- | --- | --- |
|  | All “Filtered Countries” |  |  |  | “First Wave” |  |  |  | “Second Wave” |  |  |  |
|  | Total Cases per Million |  | Total Deaths per Million |  | Total Cases per Million |  | Total Deaths per Million |  | Total Cases per Million |  | Total Deaths per Million |  |
|  | New Case | New Death | New Case | New Death | New Case | New Death | New Case | New Death | New Case | New Death | New Case | New Death |
| Strength Indicator R<sup>2</sup> | 0.136 | 0.082 | 0.318 | 0.394 | 0.298 | 0.015 | 0.703 | 0.295 | 0.114 | 0.141 | 0.263 | 0.443 |
| Strength Indicator Slope | 12396 | 5914 | 677 | 463 | 14011 | 1378 | 1465 | 422 | 11646 | 8889 | 532 | 474 |
| Persistence Indicator R<sup>2</sup> | 0.156 | 0.09 | 0.364 | 0.409 | 0.131 | 0.044 | 0.540 | 0.242 | 0.167 | 0.124 | 0.361 | 0.5 |
| Persistence Indicator Slope | 13314 | 8147 | 728 | 623 | 10518 | 3598 | 1455 | 573 | 13834 | 10443 | 612 | 630 |

**Table 5:**
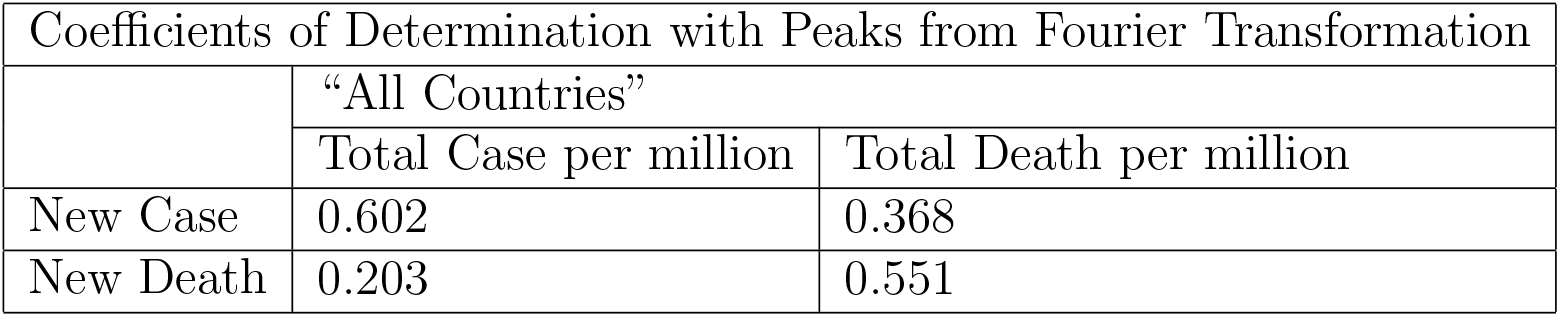
The coefficients of determination for these relationships using Fourier transformation are tabulated here.

In general, a 7-day periodicity for the daily new cases and new deaths can be confirmed using autocorrelation computation and Fourier transformation. The amplitudes of the periodicity in the autocorrelation and Fourier transformations are positively and linearly correlated with the total cases per million and total deaths per million for each country, and the coefficients of determination, or linear correlation coefficients, for most of the data are very significant - the stronger or more persistent the periodicity is, the higher the total cases and deaths per million population. The large slopes of the linear regressions show quantitatively how strong the relationships are between rising strength or persistence of the periodicity and increasing cases and deaths.

### 3.1 “All Countries” Autocorrelation

When the two categories based on the “waves” were first created, it was based mostly on a hypothesis that the countries infected later on would behave differently than the countries infected earlier because the “second wave” countries would have more information about the virus and the related prevention tactics with better preparation. The results confirm that there are some differences between the “first wave” and the “second wave” countries.

In the “first wave” countries, the new case autocorrelation was more strongly correlated with both the total cases per million and total deaths per million than with the new death autocorrelation. This suggests that within the “first wave” countries, the periodicity of the new cases is a stronger indicator of the total deaths and total cases. In particular, the new case Persistence Indicator and total death per million have a linear coefficient of determination of of 0.57 (correlation of 75%), and the new death Persistence Indicator and total death per million coefficient of determination of 0.43 (correlation of 66%), while new case Persistence Indicator and total death per million correlation of 0.29 (correlation of 54%) [Table 3].

For the “second wave” countries the linear correlations are slightly weaker and the differences across the case data categories are smaller than the corresponding ones in the “first wave” countries. There is a more direct and intuitive relationship: periodicity in new cases is more linked to total cases and periodicity in new deaths is more linked to total deaths. Looking at “second wave” countries, both new case and new death autocorrelations are more strongly correlated with total deaths per million than with total cases per million.

**Figure 5:**
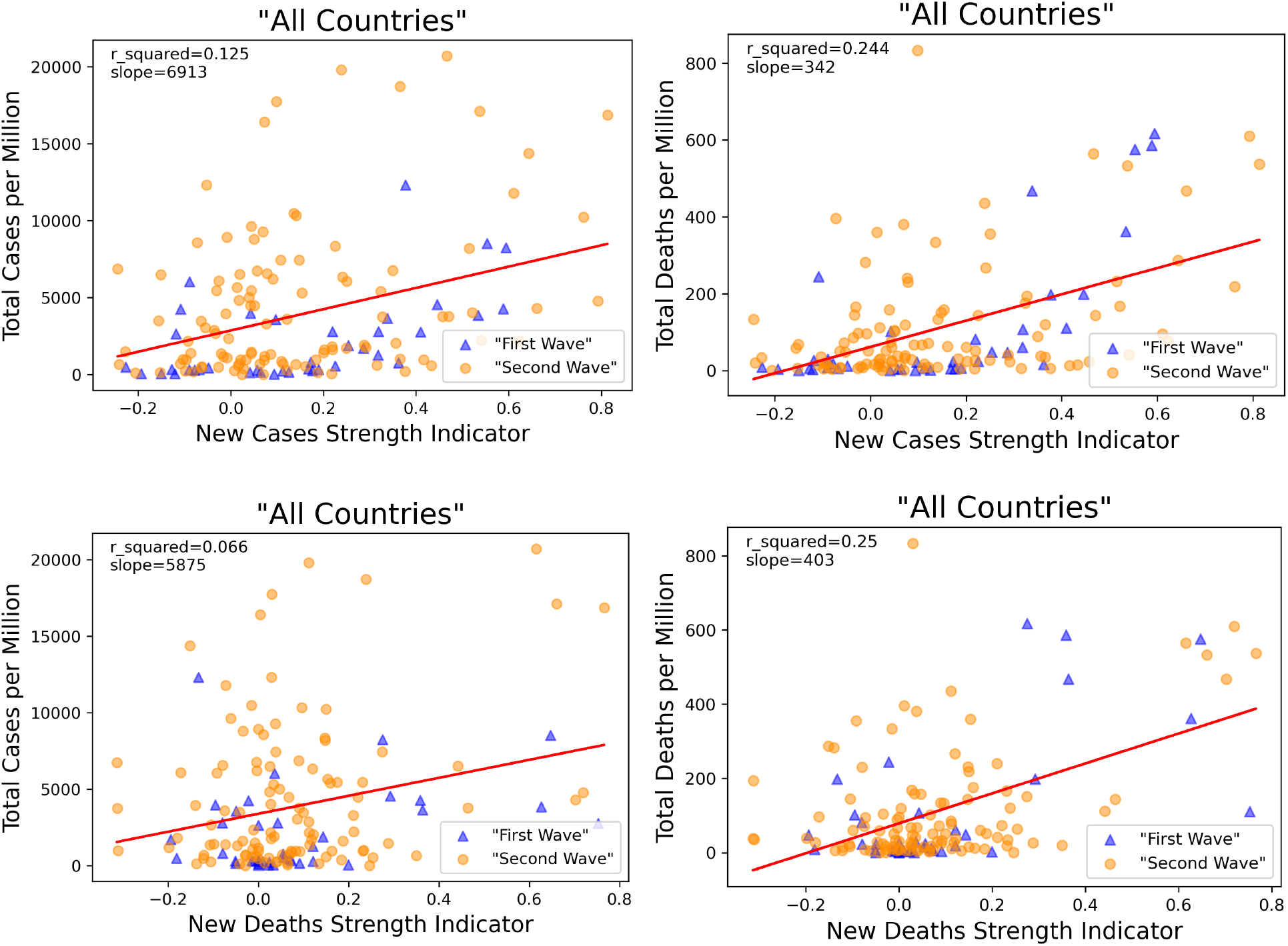
“All countries” periodicity Strength Indicators vs total cases and deaths per million.

In addition to the difference in the coefficients of determination, we can also observe that the regression line slopes corresponding to total cases per million for “first wave” countries are slightly smaller than the slopes corresponding to those in “second wave” countries. Whereas the slopes corresponding to total deaths per million in “first wave” countries are greater than in “second wave” countries. The large numerical values of the slopes show how significant the increasing periodicity is linked to the rising cases and deaths, and changing in responses leading to a reduction in periodicity could potentially lower the cases and deaths substantially.

**Figure 6:**
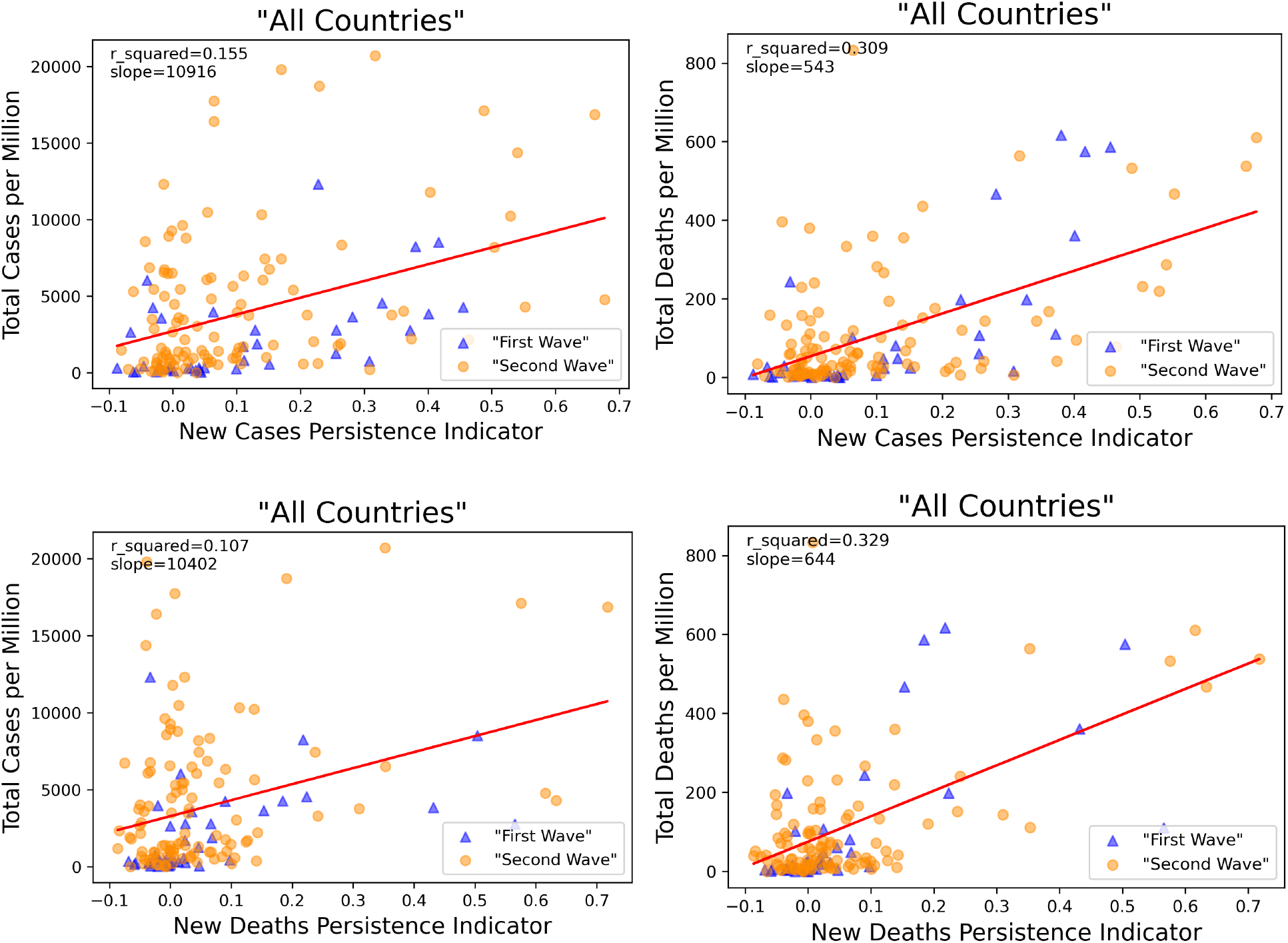
“All countries” periodicity Persistence Indicators vs total cases and deaths per million.

### 3.2 “Filtered Countries” Autocorrelation

Selecting countries with more substantial case data allows us to examine the more pronounced trends separately. The overall relationships among the coefficients of determination are the same across the “filtered countries” and “all countries”, but the range of the coefficients is much greater for the filtered countries. Looking at the autocorrelation and coefficients of determination for “filtered countries”, we see the highest coefficient of determination in the whole analysis - the coefficient of determination between the Strength Indicator for new cases in “first wave” countries’ and total deaths per million is 0.70 (linear correlation of 84%). This suggests that the strength, and not so much the persistence, of the periodicity in the new cases is more strongly related to the total deaths per million in the “first wave” countries that have significant data. Since in “first wave” countries, the situations are changing rapidly, the threats more urgent, the consequences less known, and the responses more forceful, consequently the modulations are more volatile, and the autocorrelations decay faster over multiple weeks for these countries.

**Figure 7:**
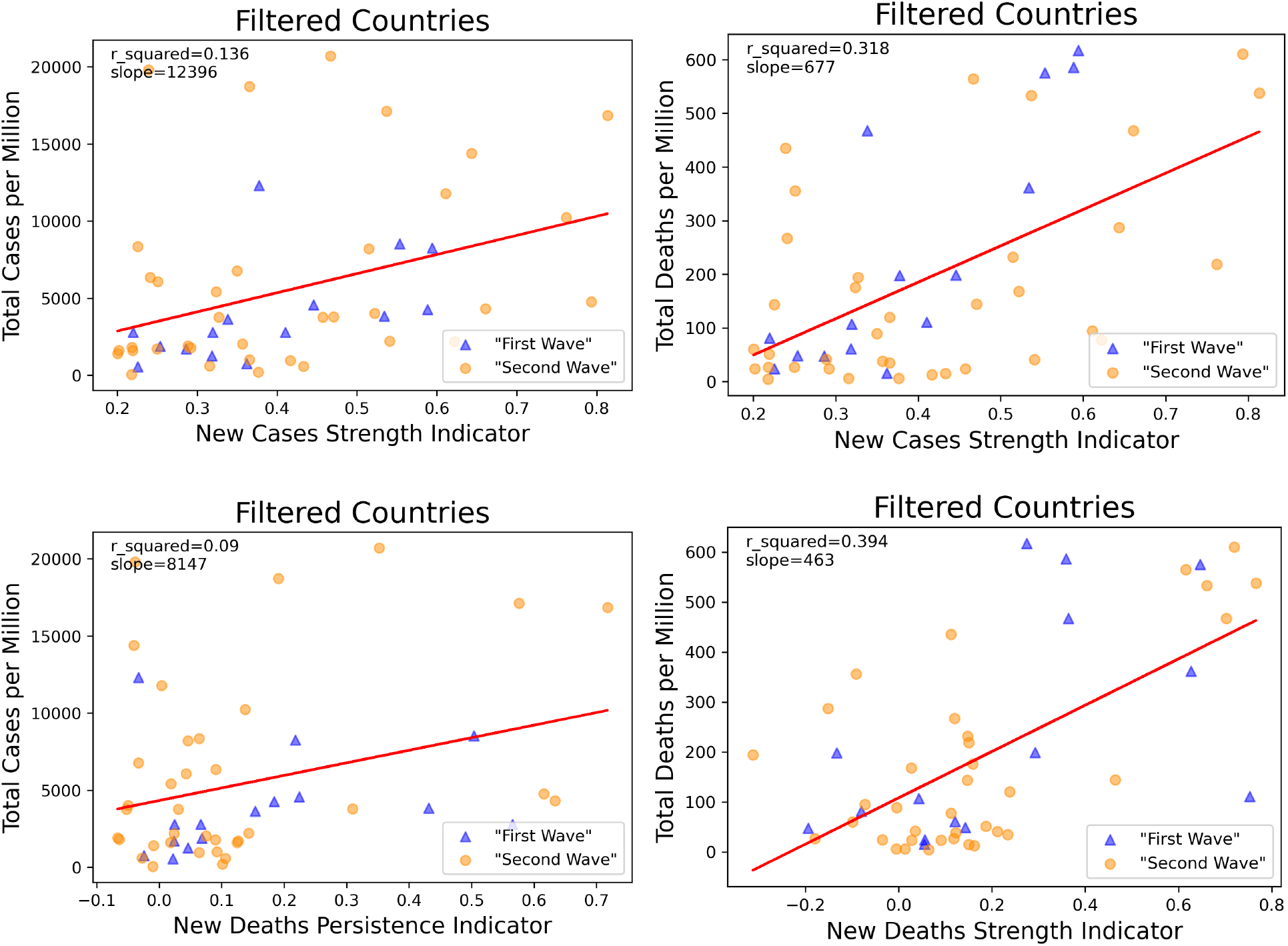
“Filtered countries” Strength Indicators vs total cases https://www.overleaf.com/project/5f692c3b0af2300001f07c8f and deaths per million.

Looking at the “filtered countries”, the relationships between the different autocorrelations and total deaths and cases per million reflect similar relationships between new cases and new deaths with the societal response and the healthcare response in the unfiltered country analysis. However, the effects are amplified, or coefficients of determination are stronger. This pattern is also reflected in the slopes of the regression lines.

**Figure 8:**
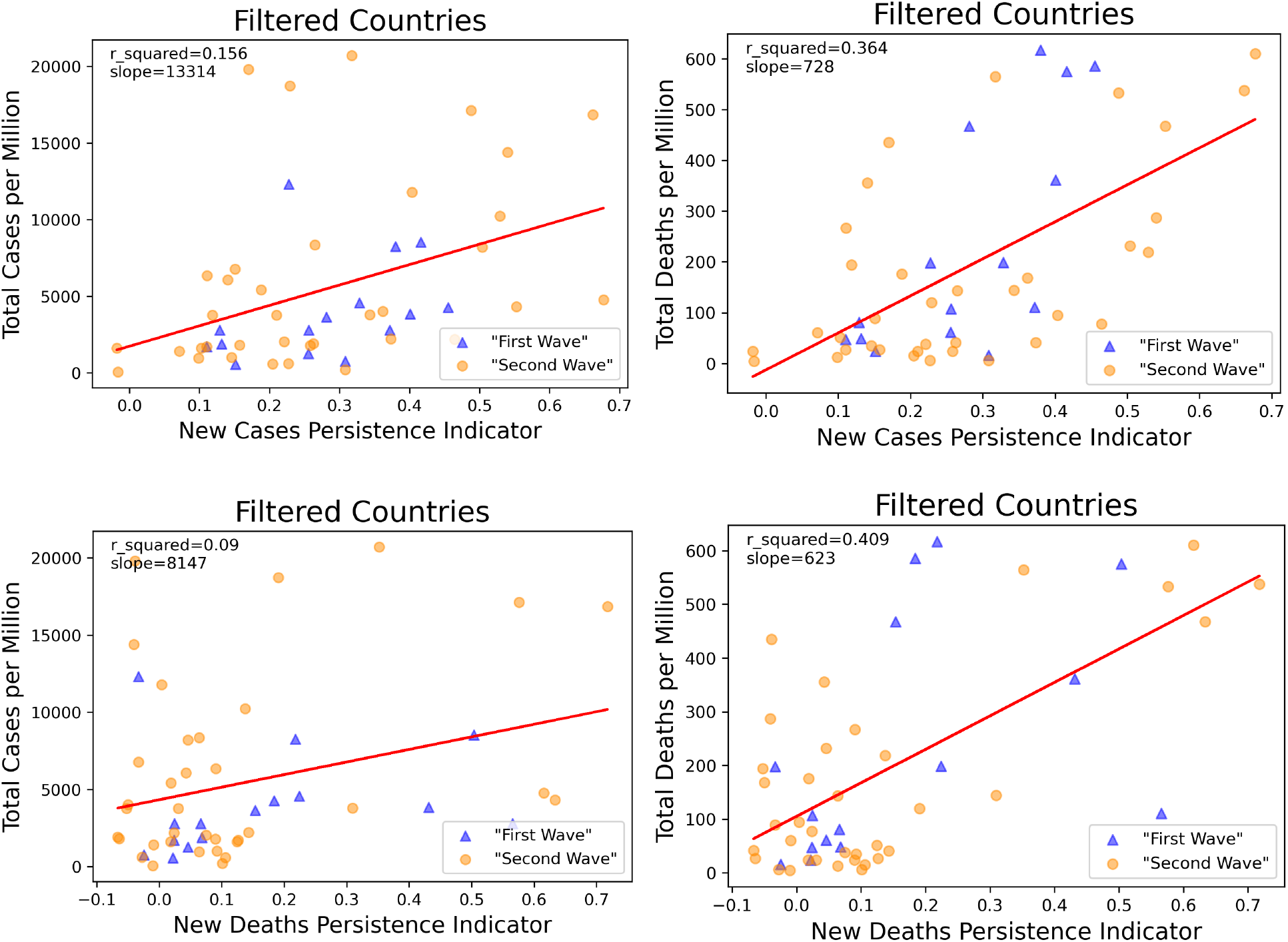
“Filtered countries” Persistence Indicators vs total cases and deaths per million.

### 3.3 Fourier Transformation

In the Fourier transformation analysis, we also see a significant linear correlation between the amplitudes of the Fourier transformation at the 1/7 frequency and total cases and deaths per million population. The results are consistent with the previous analysis[5]. A Fourier transformation can produce stronger signals if the data series are sufficiently long; the coefficients of determination with the Fourier transformation for all countries are stronger than with the autocorrelation indicators. However, autocorrelation can produce relatively significant signals early on. Having a more comprehensive understanding of the situation early and quickly can help devise more effective and timely strategies and measures to combat the pandemic in a more timely manner.

**Figure 9:**
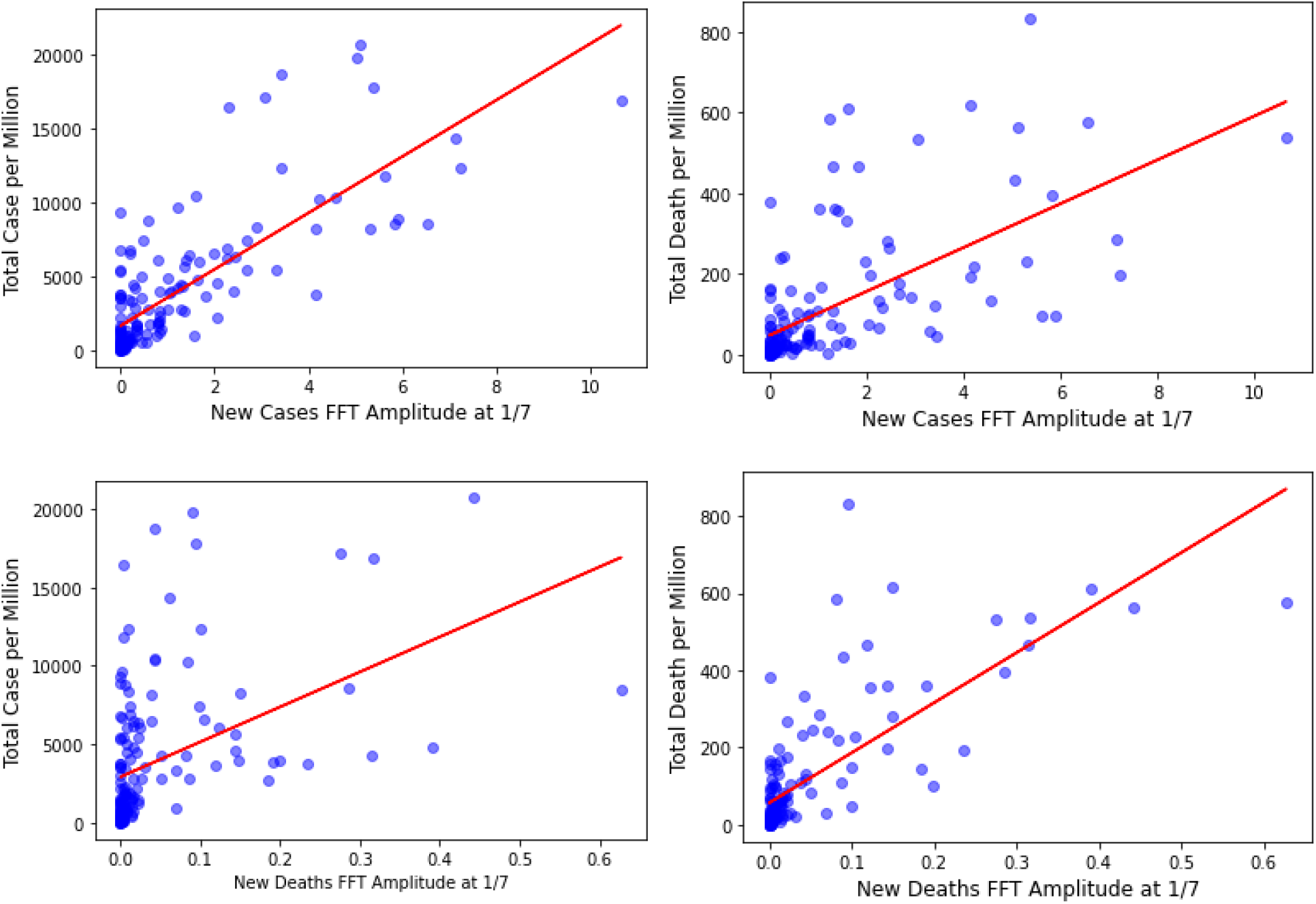
The amplitude of the Fourier transformation data at 1/7 frequency for “all countries” are shown here, plotted against the total cases and deaths per million.

## 4 Discussion

Although this study does not cover the causation or mechanism between this observed weekly periodicity and the total deaths and cases per million, these first-reported linear (cross-) correlations are significant enough to suggest the urgent need for more exploration on this subject. Understanding more about how human activities affecting the infectious disease spread is crucial in these times. We are still in the midst of the COVID-19 pandemic, and the conventionally defined second wave has already arrived, while the first wave has not even receded in other countries. The strong linear correlations appear to indicate that the societies still maintaining a normal 7-day work/rest week during this crisis have committed less overall to strict quarantine guidelines and emergency responses, resulting in higher deaths, longer shutdowns, and slower reopening and recovery.

The coefficients of determination for the linear regressions between the Persistence Indicators and case data are typically greater than those between the Strength Indicators and case data, among most categories of the analyzed countries. This suggests that the persistence of the weekly periodicity may affect the pandemic progression and outcome more. Although seemingly low, we believe that the coefficients of determination are significant enough to ascertain that periodicity is related to total deaths and total cases per million. Although the coefficients of determination is not high enough to be used as a predictive tool, it proves that the relationship between periodicity and total deaths and total cases per million is not random.

Looking at the “all country” results, the new case periodicity is likely linked more strongly to the societal response, where a weaker periodicity suggests a societal response that has focused more strongly on changing the majority’s typical behavior in the face of a pandemic.

The strong positive linear correlation might suggest that in the “first wave” countries, the stronger the societal response, such as the more strict quarantine and social distancing, and prevalent PPE use, is linked to fewer total cases and total deaths. In addition, the new death periodicity is more strongly linked to the healthcare response, where a weaker periodicity likely suggests that the healthcare system has dedicated more time, efforts, and resources in treating and caring the infected in an emergency. Whereas a stronger periodicity indicates more business-as-usual during the pandemic. The correlation here suggests that in the “first wave” countries, the stronger the healthcare responses are, such as affordably accessible emergency hospital care and availability of sufficient time, equipment, and personnel, the weaker the weekly periodicity is, and the fewer the total deaths and total cases there are. Higher periodicity in new cases and new deaths leads to more total cases and total deaths, and suggests that societal and institutional healthcare response has substantial room to improve.

The coefficients of determinism is slightly lower for the “second wave” countries. A “second wave” country likely has more experiences and information made available from the “first wave” countries regarding COVID-19, so compared to the “first wave” countries, it makes sense that the periodicity is not as strongly linked to the total cases or total deaths. This also indicates that over time, the linkage between the periodicity and pandemic progression and outcome should weaken.

Combining the observations from above and looking holistically, even with the differences between the two categories of countries, we observe that generally the strength and persistence of periodicity are more correlated to the total deaths per million. This confirms another initial hypothesis that the periodic pattern is likely more connected to the institutional healthcare response.

Exploring the reasons why a 7-day periodicity is positively linked to the total cases and deaths may provide policy makers with a new perspective to design and issue policies and implement them more effectively that would improve the outcome. Since this coronavirus has an incubation period typically shorter than 14 days, a strict temporary lockdown with strong protection and cleaning measures only needs to last a few weeks for the majority of the infectious sources to be identified, contained, or eliminated. In the past few months, individuals living in countries with significant daily new cases for months have felt the negative effects of the lagging economies and the lack of normal social interactions, but little can be done when COVID-19 is still in full swing. Our results show that if a group’s behaviour shows strong periodicity, indicating lax or uneven lockdowns or protections, not treating the pandemic seriously as a real emergency, or pre-maturely going back to business as usual, the more likely COVID-19 will cause more damages and end later. The sooner COVID-19 ends, the sooner we can all recover and thrive again.

This pandemic is still ongoing. COVID-19 has infected more than 60 million people, claiming 1.4 million lives, as of November 24th, truly making this pandemic a century defining event [26]. There is no strategizing or negotiating with a virus. If the periodicity and its effects are strong, we should understand them as much as we can and use the new found knowledge to improve our societal and institutional responses with a new perspective and corresponding measures. Having analyzed the 7-day periodicity, it is apparent that this is a valid and useful parameter and indicator that can provide further information on a developing pandemic, and give insight to the potential non-virological and non-pharmacological causes contributing to the eventual outcomes. We should use the information and knowledge learned from this analysis and change our behaviour and coordinate, plan, and implement policies and strategies to reduce the economic cost of lengthy shutdown and human toll.

### 4.1 Future Work

There is a great deal of work to do after this preliminary study, both in regards to COVID-19 specifically and infectious diseases in general. The analysis can be expanded to examine different categories instead of the two waves in this study, which can give deeper insight into the geographical, climate, and cultural variables in the spread of the infectious diseases. We can also conduct the analysis to observe countries in different time phases, and directly reveal the changes in periodicity over time as countries react to the pandemic progressively. This has implications on comprehensively understanding more specific responses, especially since this is a rapidly developing situation where organization and governments are evolving their policies, as well as finding more effective and efficient dynamic solutions to pandemic containment.

Expanding the discoveries of this study to make a tool based on a first-principle-based approach, it is possible to incorporate high-frequency periodicity into an epidemic model to simulate its effects on the pandemic progression and use it as a parameter to predict and modify the progression and eventual outcome. It is useful in better understanding the effects of periodicity and assessing the effectiveness of different policies and responses. Finally, periodicity in other infectious diseases should be further analyzed; discovering and understanding how diseases like seasonal influenza are related to the periodicity due to human activities can give public healthcare professionals and policy makers more information to prevent disease spread and save lives.

## Data Availability

The data is open-source obtained from Our World in Data.

https://covid.ourworldindata.org/

## Acknowledgements

This paper and the research behind it would be impossible to write without the support from my family and friends.

I want to thank Dr. Ning Li for his mentorship on conducting research and writing a technical paper. Dr. Li and Dr. Robert Ecke first communicated on the observation of the apparent weekly periodicity in the reported case data in May, 2020, which contributed to the start of my research project in June.

I have had the privilege of discussing my preliminary work and results with Dr. Ruian Ke an Dr. Paul Li who have also given me valuable suggestions and advice and this paper. In addition, I am thankful for Professor Bernard de Jong who helped me with technical writing and an alternative perspective on this topic.

## Supplementary Materials

These materials are supplied for more comprehensive and in-depth assessments of the relationships between the periodicity indicators and the progression and outcomes of the pandemic for the various categories of countries.

**Figure S1:**
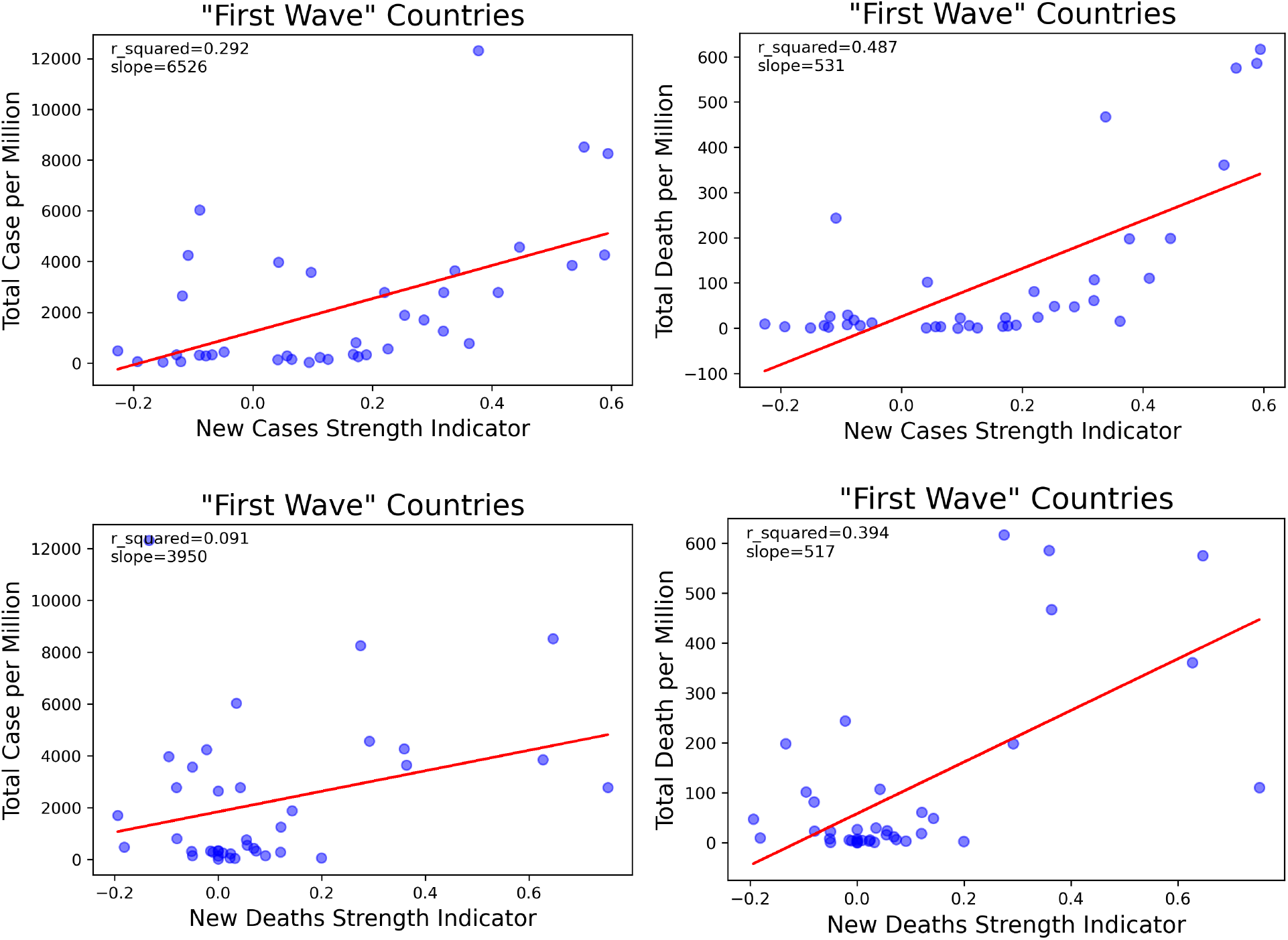
The Strength Indicators vs the total cases and deaths per million population for the “first wave” countries.

**Figure S2:**
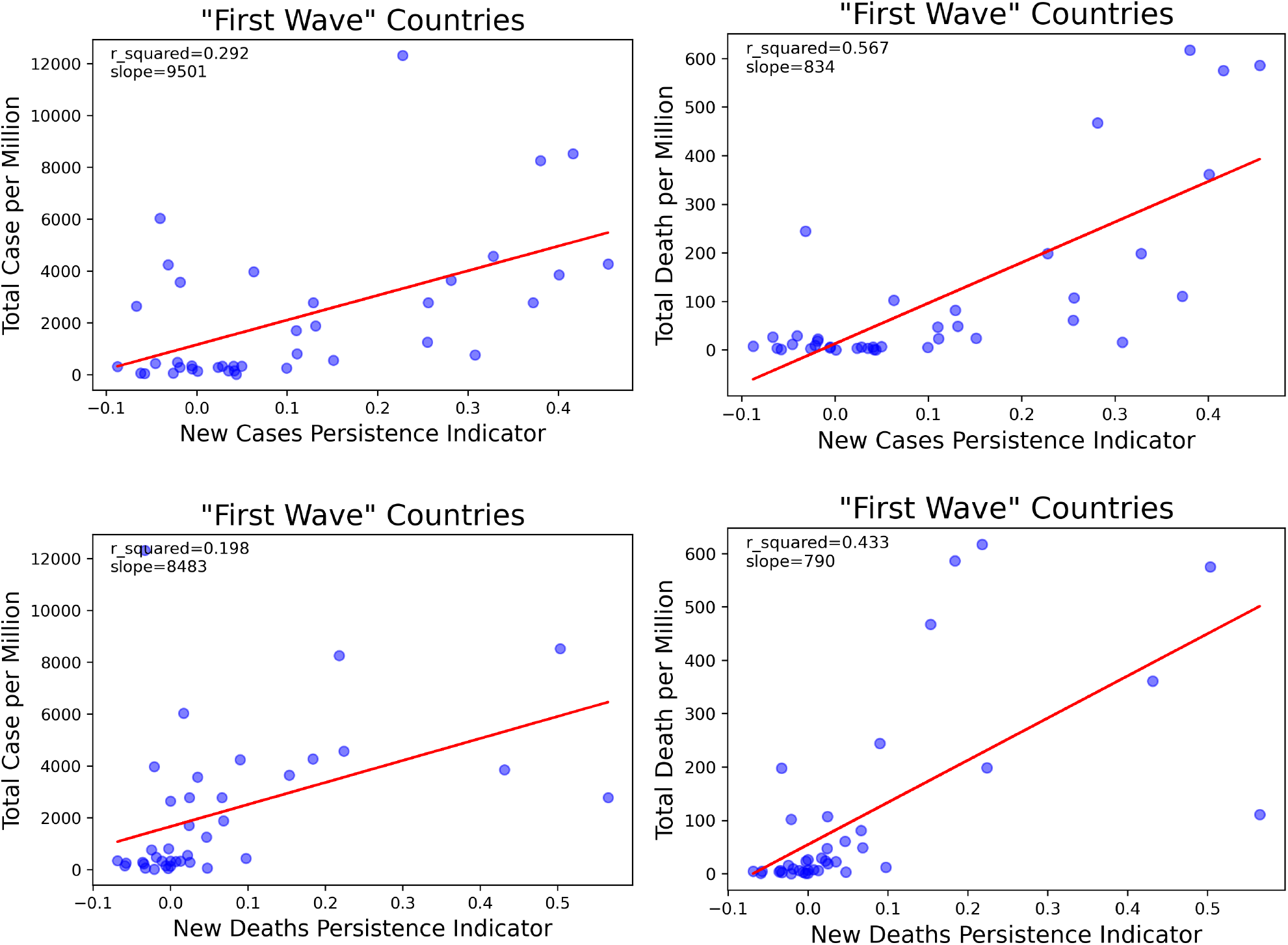
The Persistence Indicators vs the total cases and deaths per million population for the “first wave” countries.

**Figure S3:**
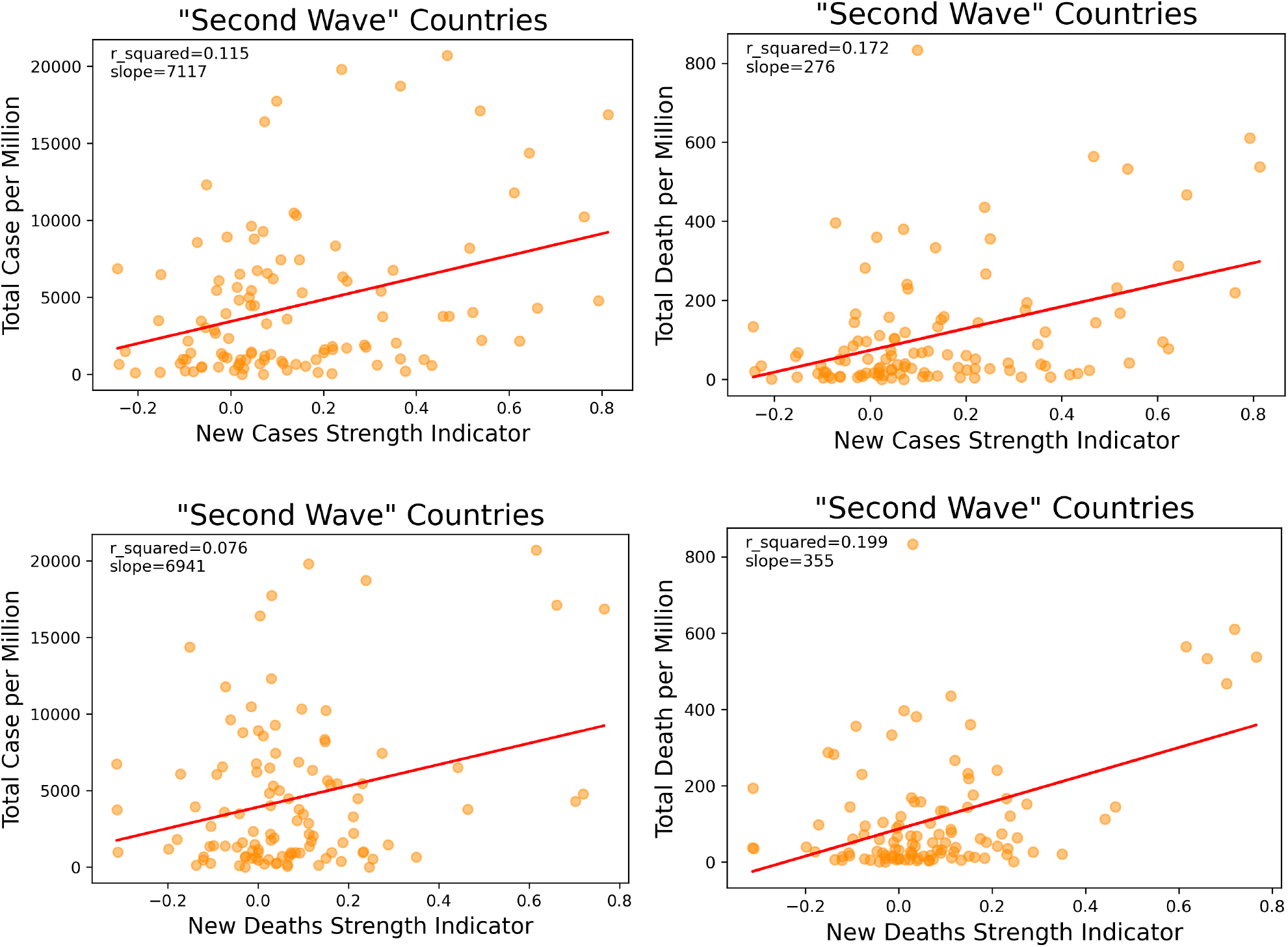
The Strength Indicators vs the total cases and deaths per million populations for the “second wave” countries.

**Figure S4:**
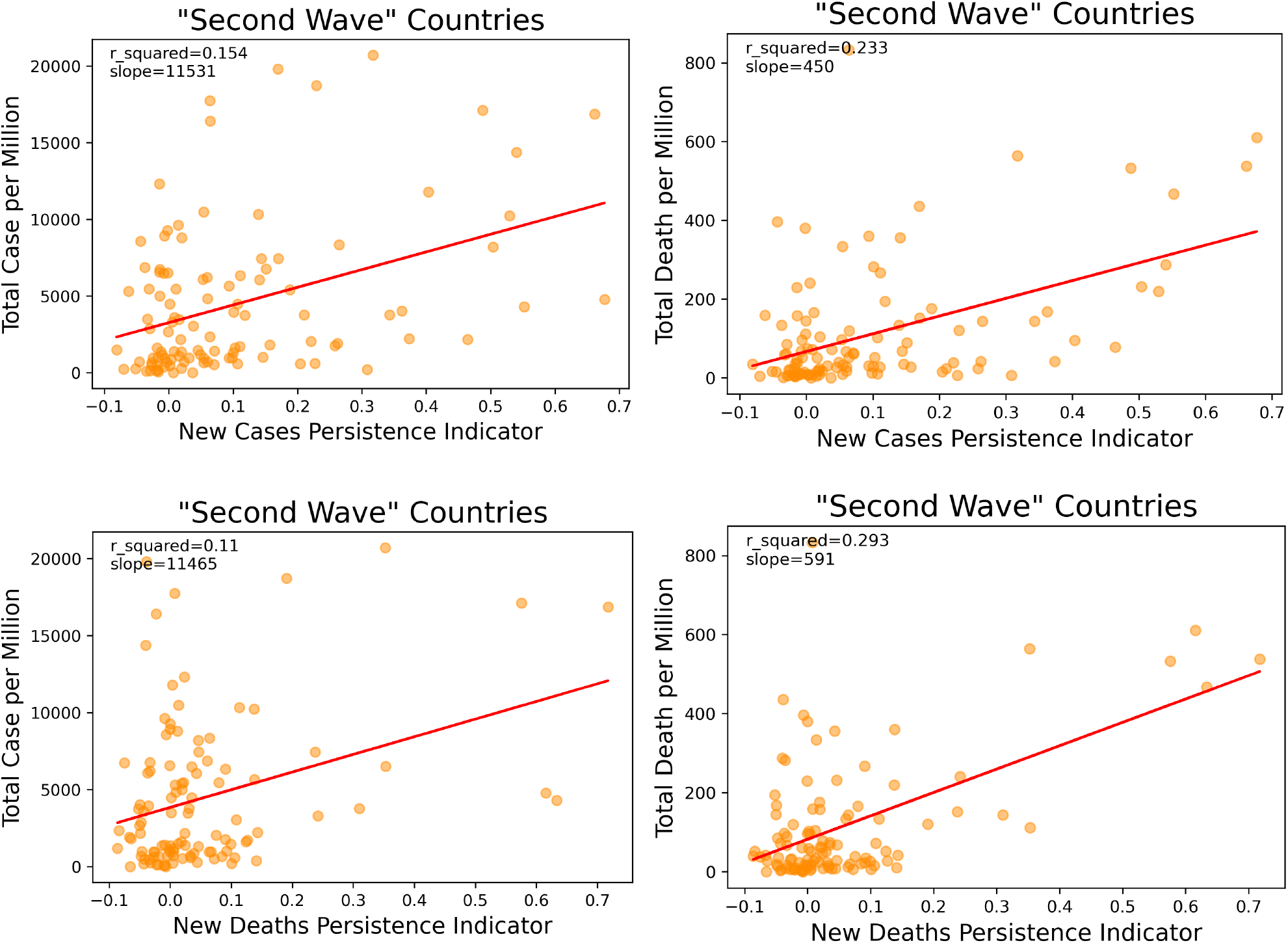
The Persistence Indicators vs the total cases and deaths per million populations for the “second wave” countries.

**Figure S5:**
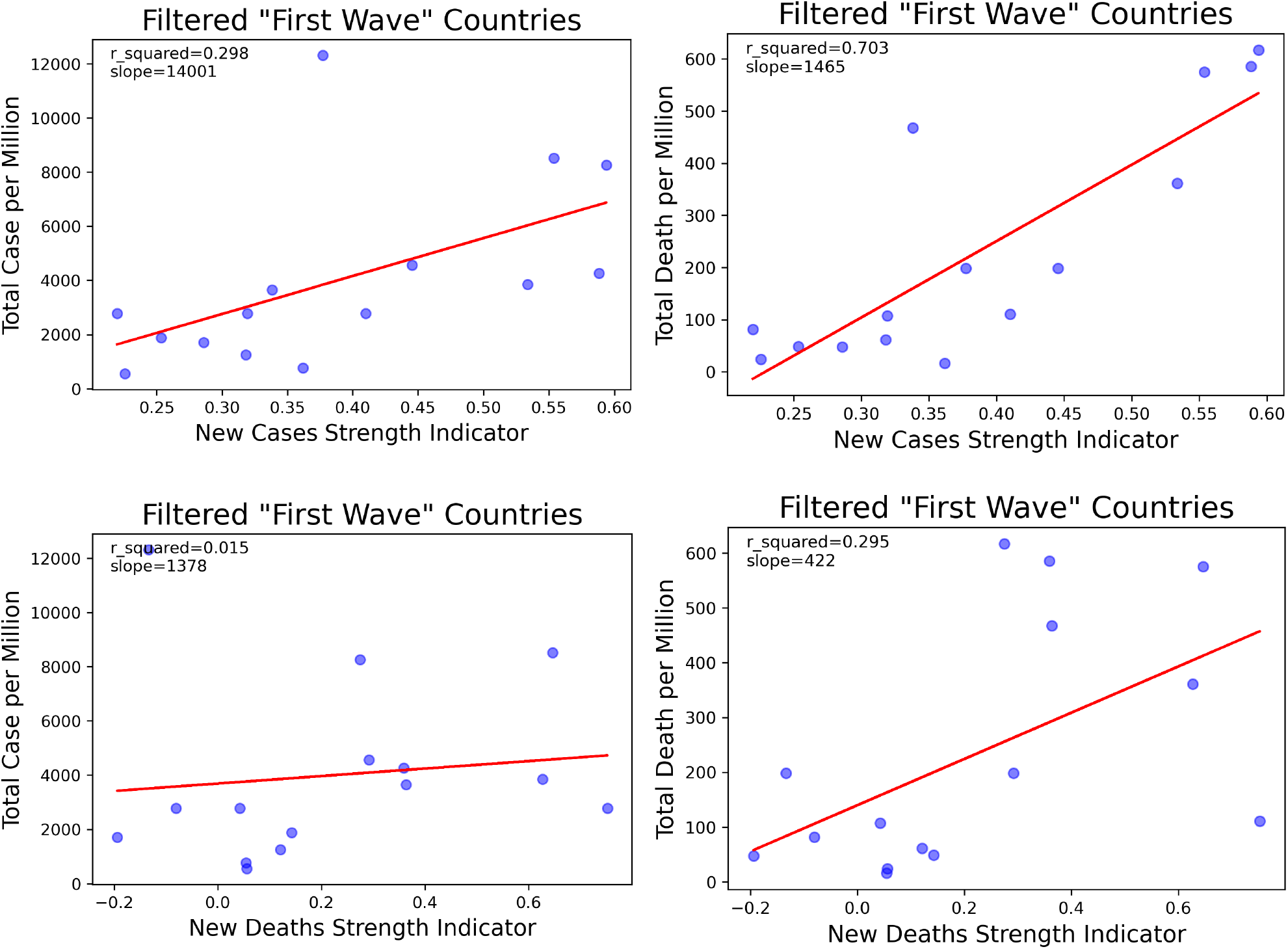
The Strength Indicators vs the total cases and deaths per millions populations for the filtered “first wave” countries.

**Figure S6:**
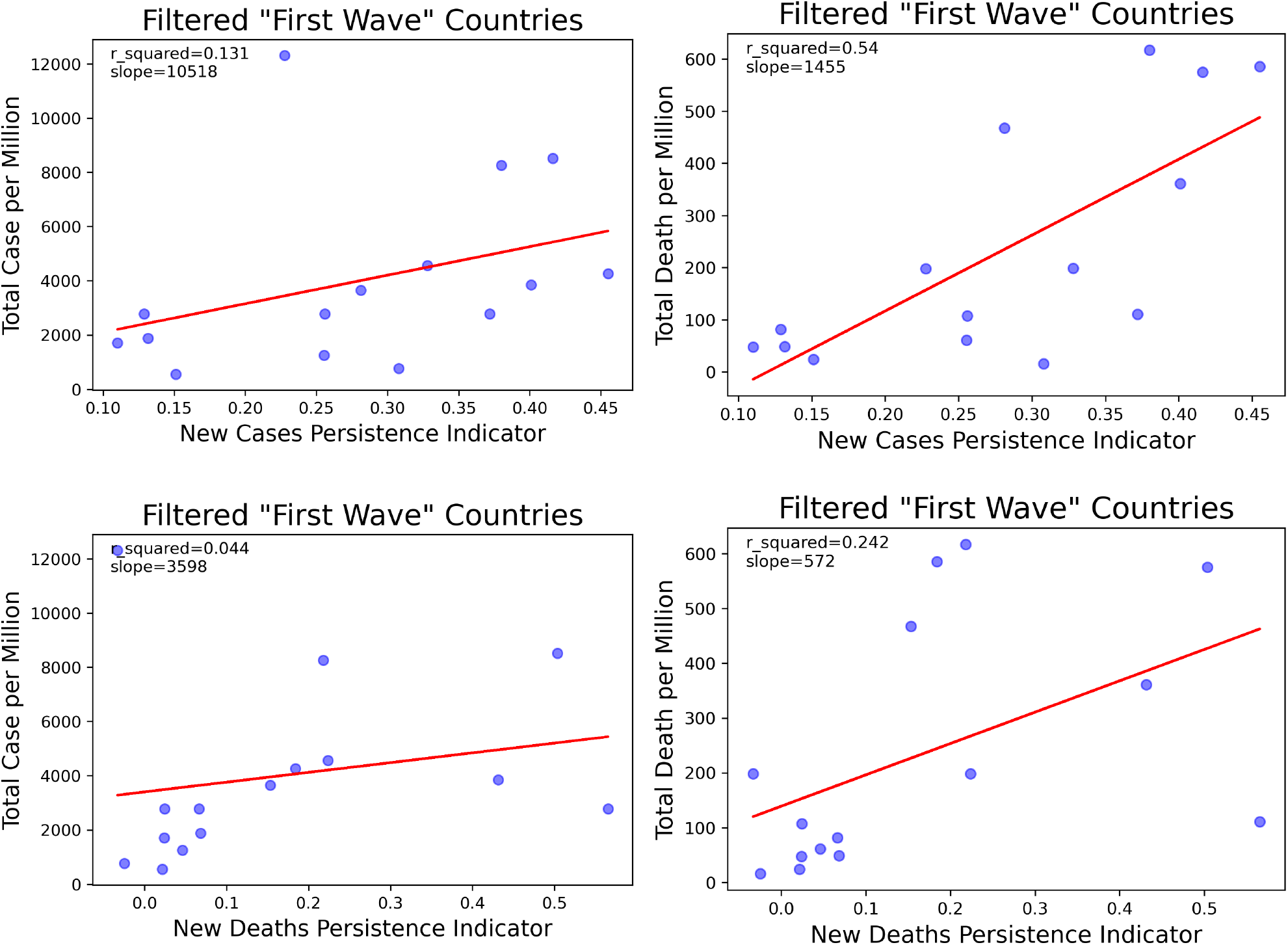
The Persistence Indicators vs the total cases and deaths per million population for the filtered “first wave” countries.

**Figure S7:**
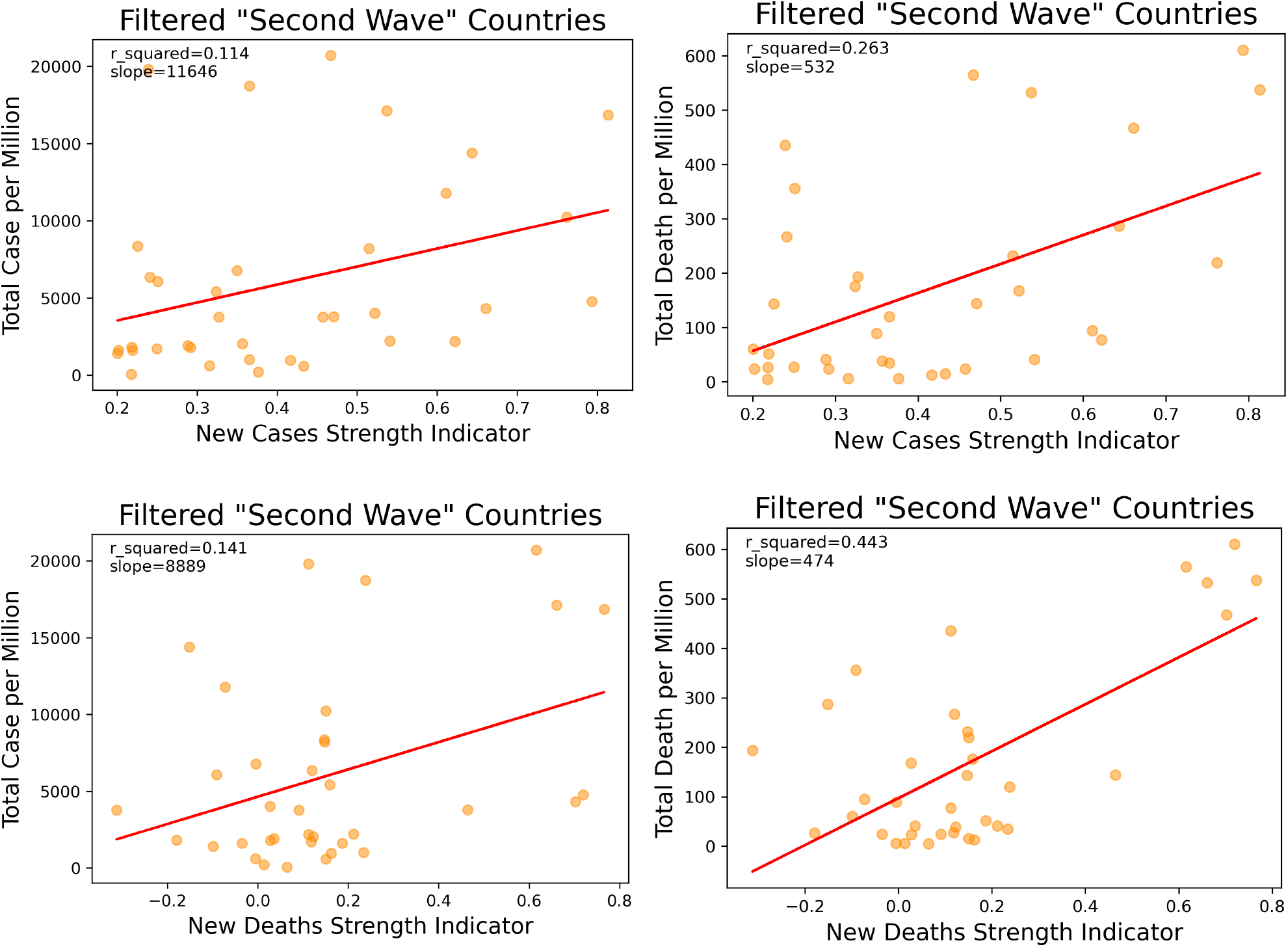
The Strength Indicators vs the total cases and deaths per million population for the filtered “second wave” countries.

**Figure S8:**
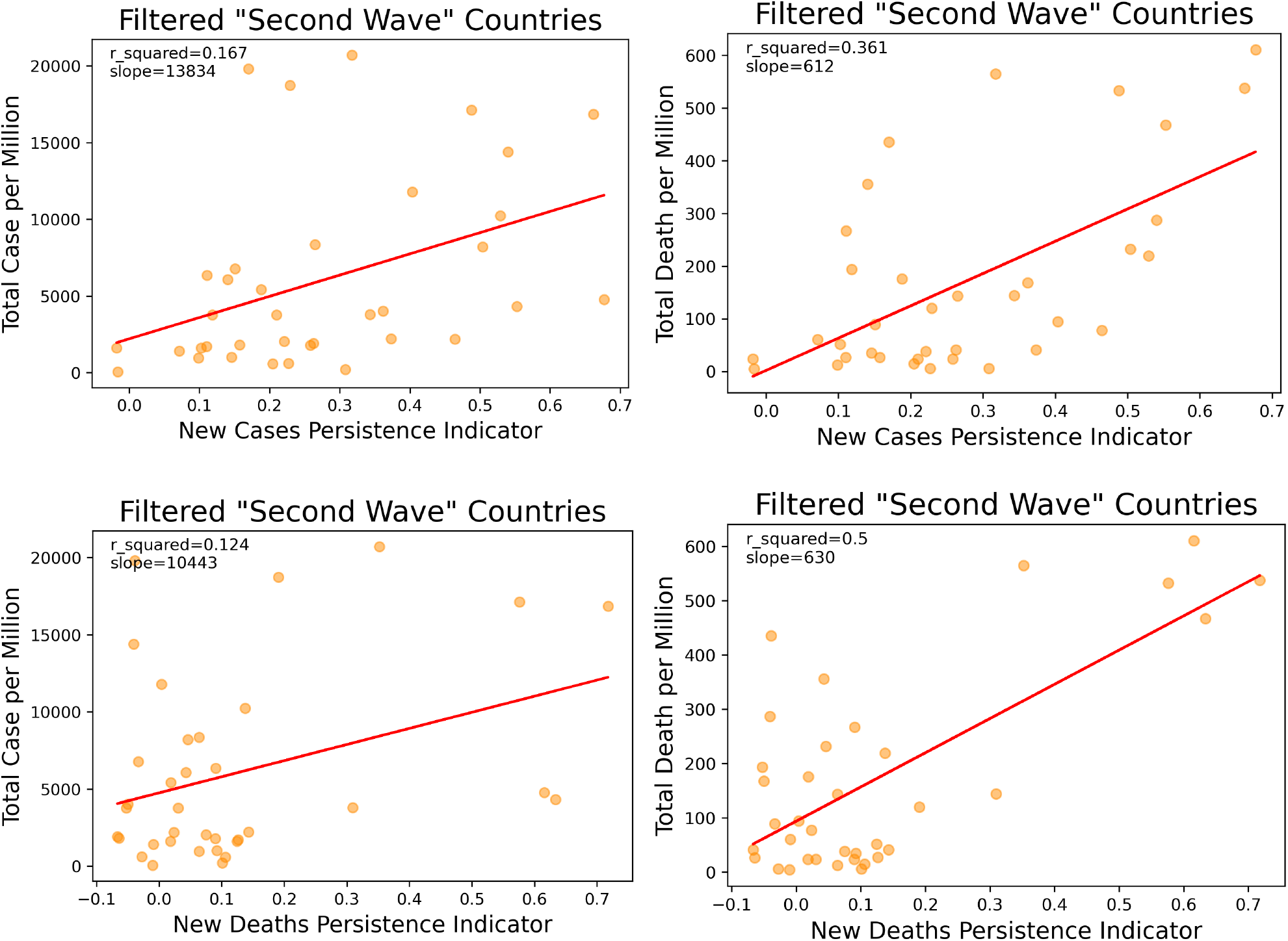
The Persistence Indicators vs the total cases and deaths per million population for the filtered “second wave” countries.

## References

[1] Worldometers.info, “Countries where COVID-19 has spread.” https://www.worldometers.info/coronavirus/countries-where-coronavirus-has-spread/., 2020 (Accessed September 13, 2020).

[2] N. Li, “Contracting economies can potentially kill more - tough road to recoveries and hard balance of risks.” https://medium.com/@powertocw/contracting-economies-can-potentially-kill-more-tough-road-to-recoveries-and-hard-balance-of-9ed6c4885c4c, Apr 2020 (Accessed September 17, 2020).

[3] N. Li, “Are stock markets voting or weighing pandemic responses and impacts.”https://medium.com/@powertocw/are-stock-markets-voting-or-weighing-pandemic-responses-and-impacts-f64025f06006, July 2020 (Accessed September 30, 2020).

[4] https://raw.githubusercontent.com/owid/covid-19-data/master/public/data/owid-covid-data.csv, 2020 (Accessed August 23, 2020.

[5] J. D. Muccigrosso, “Weekly periodicity in Italy’s COVID-19 tests & new cases.” https://jmuccigr.github.io/blog/2020/04/10/Weekly-Periodicity/, Apr 2020 (Accessed September 15, 2020).

[6] C. Unnikrishnan, “Globally coherent weekly periodicity in the COVID-19 pandemic,” medRxiv, 2020.

[7] T. Pavlicek, P. Rehak, and P. Kral, “Oscillatory dynamics in infectivity and death rates of COVID-19,” medRxiv, 2020.

[8] B. Wilder, M. Charpignon, J. A. Killian, H.-C. Ou, A. Mate, S. Jabbari, A. Perrault, A. Desai, M. Tambe, and M. S. Majumder, “The role of age distribution and family structure on COVID-19 dynamics: A preliminary modeling assessment for Hubei and Lombardy,” Available at SSRN 3564800, 2020.

[9] R. Li, S. Pei, B. Chen, et al., “Substantial undocumented infection facilitates the rapid dissemination of novel coronavirus (SARS-CoV-2)[published online ahead of print March 16, 2020],” Science, vol.10, 2020.

[10] C. Anastassopoulou, L. Russo, A. Tsakris, and C. Siettos, “Data-based analysis, modelling and forecasting of the COVID-19 outbreak,” PloS one, vol. 15, no. 3, p. e0230405, 2020.

[11] G. C. Calafiore, C. Novara, and C. Possieri, “A modified sir model for the COVID-19 contagion in Italy,” arXiv preprint 2003.14391, 2020.

[12] Z. Liu, p. Magal, O. Seydi, and G. Webb, “A COVID-19 epidemic model with latency period,” Infectious Disease Modelling, vol. 5, pp. 323–337, 2020.

[13] L. Peng, W. Yang, D. Zhang, C. Zhuge, and L. Hong, “Epidemic analysis of COVID-19 in China by dynamical modeling,” arXiv preprint 2002.06563, 2020.

[14] A. Mollalo, B. Vahedi, and K. M. Rivera, “Gis-based spatial modeling of COVID-19 incidence rate in the continental United States,” Science of The Total Environment, p. 138884, 2020.

[15] K. Chatterjee, K. Chatterjee, A. Kumar, and S. Shankar, “Healthcare impact of COVID-19 epidemic in India: A stochastic mathematical model,” Medical Journal Armed Forces India, 2020.

[16] M. Y. Li, W. Liu, C. Shan, and Y. Yi, “Turning points and relaxation oscillation cycles in simple epidemic models,” SIAM Journal on Applied Mathematics, vol. 76, no. 2, pp. 663–687, 2016.

[17] M. Alexander and S. Moghadas, “Periodicity in an epidemic model with a generalized non-linear incidence,” Mathematical Biosciences, vol. 189, no. 1, pp. 75–96, 2004.

[18] S. M. Kissler, C. Tedijanto, E. Goldstein, Y. H. Grad, and M. Lipsitch, “Projecting the transmission dynamics of SARS-CoV-2 through the postpandemic period,” Science, vol. 368, no. 6493, pp. 860–868, 2020.

[19] Worldometers.info, “Daily new cases in the United States.” https://www. worldometers.info/coronavirus/country/us/, September 2020 (Accessed September 23, 2020).

[20] “How infections spread.” https://www.cdc.gov/infectioncontrol/spread/index.html., Jan 2016 (Accessed September 15, 2020).

[21] P. Virtanen, R. Gommers, T. E. Oliphant, M. Haberland, T. Reddy, D. Cournapeau, E. Burovski, p. Peterson, W. Weckesser, J. Bright, S. J. van der Walt, M. Brett, J. Wilson, K. J. Millman, N. Mayorov, A. R. J. Nelson, E. Jones, R. Kern, E. Larson, C. J. Carey, İ. Polat, Y. Feng, E. W. Moore, J. VanderPlas, D. Laxalde, J. Perktold, R. Cim-rman, I. Henriksen, E. A. Quintero, C. R. Harris, A. M. Archibald, A. H. Ribeiro, F. Pedregosa, p. van Mulbregt, and SciPy 1.0 Contributors, “SciPy 1.0: Fundamental Algorithms for Scientific Computing in Python,” Nature Methods, vol. 17, pp. 261–272, 2020.

[22] T. E. Oliphant, A guide to NumPy, vol. 1. Trelgol Publishing USA, 2006.

[23] J. D. Hunter, “Matplotlib: A 2d graphics environment,” Computing in science & engineering, vol. 9, no. 3, pp. 90–95, 2007.

[24] Worldometers.info, “Coronavirus cases.” https://www.worldometers.info/ coronavirus/, 2020 (Accessed September 15, 2020).

[25] “Autocorrelation.” https://www.statisticssolutions.com/autocorrelation/, (Accessed September 15, 2020).

[26] “Coronavirus cases.”

